# The Association between Poverty and Gene Expression within Peripheral Blood Mononuclear Cells in a Diverse Baltimore City Cohort

**DOI:** 10.1101/2020.05.13.20100818

**Authors:** Nicole S. Arnold, Nicole Noren Hooten, Yongqing Zhang, Elin Lehrmann, William Wood, Wendy Camejo Nunez, Roland J. Thorpe, Michele K. Evans, Douglas F. Dluzen

**Affiliations:** Department of Biology, Morgan State University, Baltimore, MD; Laboratory of Epidemiology and Population Science, National Institute on Aging, National Institutes of Health, Baltimore, MD; Laboratory of Genetics and Genomics, National Institute on Aging, National Institutes of Health, Baltimore, MD; Program for Research on Men’s Health, Hopkins Center for Health Disparities Solutions, Johns Hopkins University, Baltimore, MD

**Author notes:** Correspondence: Douglas F. Dluzen, PhD.

## Abstract

Socioeconomic status (SES), living in poverty, and other social determinants of health contribute to health disparities in the United States. African American (AA) men living below poverty in Baltimore City have a higher incidence of mortality when compared to either white males or AA females living below poverty. Previous studies in our laboratory and elsewhere suggest that environmental conditions are associated with differential gene expression (DGE) patterns in peripheral blood mononuclear cells (PBMCs). DGE have also been associated with hypertension and cardiovascular disease (CVD) and correlate with race and sex. However, no studies have investigated how poverty status associates with DGE between male and female AAs and whites living in Baltimore City. We examined DGE in 52 AA and white participants of the Healthy Aging in Neighborhoods of Diversity across the Life Span (HANDLS) cohort, who were living above or below 125% of the 2004 federal poverty line at time of sample collection. We performed a microarray to assess DGE patterns in PBMCs from these participants. AA males and females living in poverty had the most genes differentially-expressed compared with above poverty controls. Gene ontology (GO) analysis identified unique and overlapping pathways related to the endosome, single-stranded RNA binding, long-chain fatty-acyl-CoA biosynthesis, toll-like receptor signaling, and others within AA males and females living in poverty and compared with their above poverty controls. We performed RT-qPCR to validate top differentially-expressed genes in AA males. We found that KLF6, DUSP2, RBM34, and CD19 43 are expressed at significantly lower levels in AA males in poverty and KCTD12 is higher compared to above poverty controls. This study serves as an additional link to better understand 45 the gene expression response in peripheral blood mononuclear cells in those living in poverty.

## Introduction

In 2018, 38.1 million people living in the United States (11.8% of the US population) were living in poverty. African American (AAs)in the US had the highest poverty rate of any racial demographic (20.8%; 8.9 million) (1). This compares with 15.7% (15.7 million) of whites 50 living in poverty, whom have the lowest poverty rate in the US (1). Black household median income levels in the US were just 59% of the median household income levels of non-Hispanic Whites; whereas Hispanics were 73% of non-Hispanic Whites and Asian households were 123% (1).

The influence of poverty on health and lifespan is associated with both chronic exposure to poverty as well as sudden wealth loss (2,3). Living in poverty or low socioeconomic status (SES) also increases the risk of disability and all-cause mortality in the US and around the world (4–7). These associations can be due to a variety of geopolitical factors and other social determinants of health. For example, neighborhoods with social or economic disadvantages such as elevated crime and violence levels can stagnant physical activity in local residents (8,9 9) and neighborhood segregation and reduced walkability and greenspace are associated with increased risk for cardiovascular diseases (CVDs), particularly within minority populations (10–12). Food insecurity can also predispose individuals to chronic cardiometabolic diseases such as obesity, diabetes, and hypertension (13,14).

Living in poverty and low SES increases the risk of myocardial infarction (15,16),having higher blood pressure and body mass index (17–19),reduced neuroanatomic health in white matter integrity (20) and hippocampal volume (21), and development of diabetes mellitus (22,23). AAs living in poverty are at an increased risk for early mortality, particularly among AA men (24,25). AA women in low SES living in the urban and rural south of the US were over twice as likely to have worse cardiovascular health compared with white women (18). Additionally, there is a disproportionately high incidence and prevalence of CVD and poverty-related health complications in AA men and women compared with non-Hispanic whites and other ethnicities (24–28).

While considerable work has been spent identifying the environmental risk factors associated with living in poverty, there still remains the challenge of fully understanding how these risk factors translate biologically into disease and health disparities. Biomarkers have brought some clarity into physiological changes associated with SES. For example, adults with early lifetime exposure to poverty and low SES had increased production of cortisol and greater stress-response production of the pro-inflammatory cytokine interleukin-6 (IL-6) and longer duration of IL-6 elevation in circulation (29–31).

Social determinants of health, including racial discrimination (32) and experiencing traumatic stress (33), can transcriptionally activate inflammatory pathways in blood cells. Transcriptional activation or repression of inflammatory genes and related pathways has also been linked with low SES. The whole-blood transcriptional profiles of low SES AAs showed over-expressed inflammatory pathways including those related to interleukin-8 signaling and transcription factor NF-kB signaling (34). AAs experiencing racial discrimination also had elevated expression of NF-kB, AP-1, CREB, and the glucocorticoid receptor compared with European Americans, suggesting stress-response pathways may influence the predisposition to inflammation and related disparate health conditions (32). Several of these genes are part of the conserved transcriptional response to adversity (CTRA) gene network. The CRTA is a stress response pathway mediated by the sympathetic nervous system and beta-adrenergic signaling to induce transcriptional activation of inflammatory genes in the immune system (32,35). In a recent study of over 1,000 diverse and young adults, family poverty status was associated with transcription of interferon-response factors and immune cell activation of dendritic cells and may contribute to lifespan-related predisposition to inflammatory diseases and conditions (36).

In 2018, nearly 26.1% of black or AAs were living below the federal poverty line in Baltimore City in Maryland, with individuals earning less than $12,140, and 50% of all individuals living in poverty in Baltimore City had incomes below 50% of the federal poverty line, earning approximately $6,000 per individual (37). Those living in poverty in Baltimore City, particularly AA men, have the highest risk for mortality when compared with other groups influenced by poverty, and this was linked with neighborhood income inequality (24, 25).Baltimore City is also the only county on the eastern seaboard north of Washington D.C. to have an average life expectancy at birth below 72 year of age (38).

Understanding the biological mechanisms linked with poverty status will help us elucidate how disparate health conditions arise and influence treatment outcomes, particularly in Baltimore City. In this study, we analyzed gene expression within a diverse sub cohort of AA and white, male and female individuals from the Healthy Aging in Neighborhoods of Diversity Across the Lifespan (HANDLS) study living above or below the federal poverty line (28). The objective of this study was to gain insight into the association between poverty status and differential gene expression (DGE).

## RESULTS

### Analysis of Gene Expression in Individuals Living in Poverty

In order to identify DGE patterns in individuals living in poverty, we performed a mRNA microarray of PBMCs isolated from a sub cohort of the HANDLS study (28). This sub-cohort was comprised of 54 AAs and whites (W), who were either male (M) or female (F), and self-reported living either below (BL) or above (AB) 125% of the 2004 Federal poverty guideline in Baltimore City at the time of PBMC collection (n=6-7/group; see S1 Appendix). There was no significant difference in age, cholesterol or CRP levels, monocyte or white blood cell counts, or history of diabetes, diagnosis of hypertension, or current smoking status between each of the 8 groups. After an initial data quality analysis of our microarray raw probe signals, we removed one white male and one white female living below poverty from our analysis for a total N of 52.

We used iPathway Guide software (39–42) to identify significantly and differentially-expressed genes between groups in our cohort. Genes were considered to be significant if the |logFC| > 0.60 and they had an adjusted false discovery rate (FDR) < 0.30. We identified 1,058 unique mRNAs as significant in our analysis and dependent upon the comparison being performed. There were 127 genes significantly different between AA men living below poverty compared with AA men living above (Figure 1A). Of these, seven mRNAs were up-regulated in AA men living in poverty (DUSP2, HLA-DQB1, CD19, MCOLN2, RMBM38, DDX39, and KLF6) and the remaining 120 were down-regulated (see. S2 Appendix for complete list).

**Figure 1:**
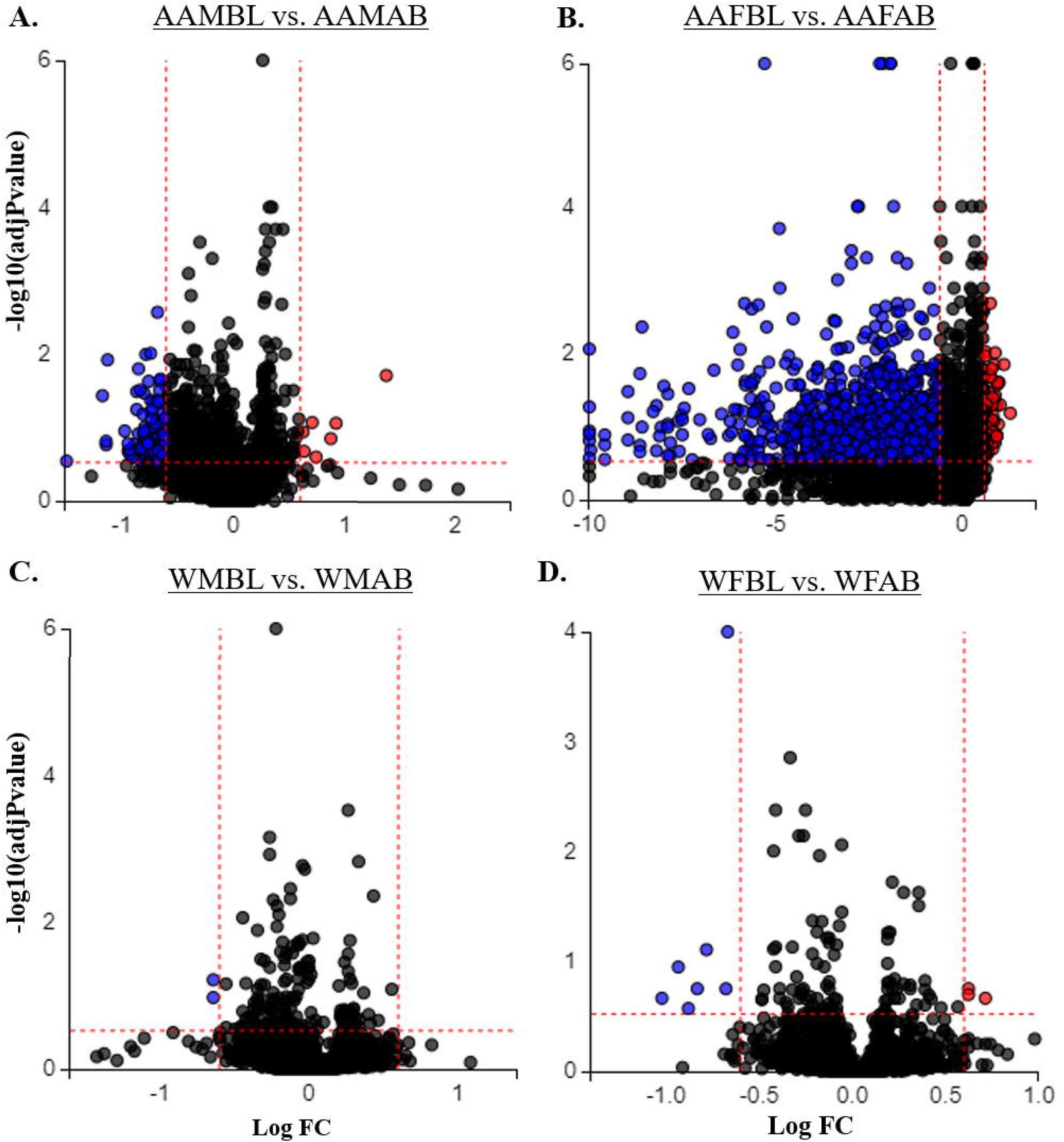
Gene expression changes associated with poverty in the HANDLS cohort. Significantly and differentially-expressed genes were identified in Baltimore City residents living below (BL) vs above (AB) poverty via microarray. Volcano plots generated in iPathway Guide show134 significantly (|LogFC| >0.6, FDR AdjPvalue < 0.3) up-regulated (red) or down-regulated (blue) genes in individuals living in poverty who were (A) African American males (AAM), (B) African American females (AAF), (C) white males (WM), or (D) white females (WF). N= 6/7 per group; see S1 Appendix for completely demographics.

We identified 993 transcripts that were significantly different between AA females living below poverty compared with AA females living above (Figure 1B). Of these transcripts, 70 genes were up-regulated and 923 were down-regulated in those living in poverty. Within whites in Baltimore, there were fewer significant genes when comparing those below poverty compared with above. We identified only two mRNAs (*MT1E and SSR4*) significantly different between white males living in poverty compared with white males living above, of which both transcripts were down-regulated (Figure 1C). When comparing white females living in poverty with those above poverty, there were 10 transcripts significantly different, three of which were up-regulated (*CD68, FBP1, and FN1*) and 7 down-regulated (*EBF, TMEM16J, MCOLN2, AIM2, BLK, FCRL5, and HLA-DQB1*; Figure 1D).Table 1 lists the top 10 significantly up-regulated and down-regulated genes.

We next sought to identify the overlap of significant genes between AA males (AAMs),AA females (AAFs), white females (WFs), and white males (WMs) living in poverty in order to identify common and unique mRNA profiles associated with poverty in each group. We found that a majority of the 1,058 significant transcripts were unique to each demographic group (Figure 2A; see. S2 Appendix for complete list). AAFs living in poverty had 921 transcripts which were significantly different with above poverty controls. AAMs living in poverty had 54 genes significantly different with AAM living above poverty. Additioanlly, there were 71 signficant transcripts that were shared between AA men and AA women living in poverty. WMs living in poverty had only two significantly different trancripts compared with WMs living above poverty (MT1E and SSR4). We observed that AAMs shared two significant transcripts with WFs (HLA-DQB1 and MCOLN2) and AAFs shared one (early B cell factor 1, EBF, Figure 2A) with WFs.

**Table 1.**
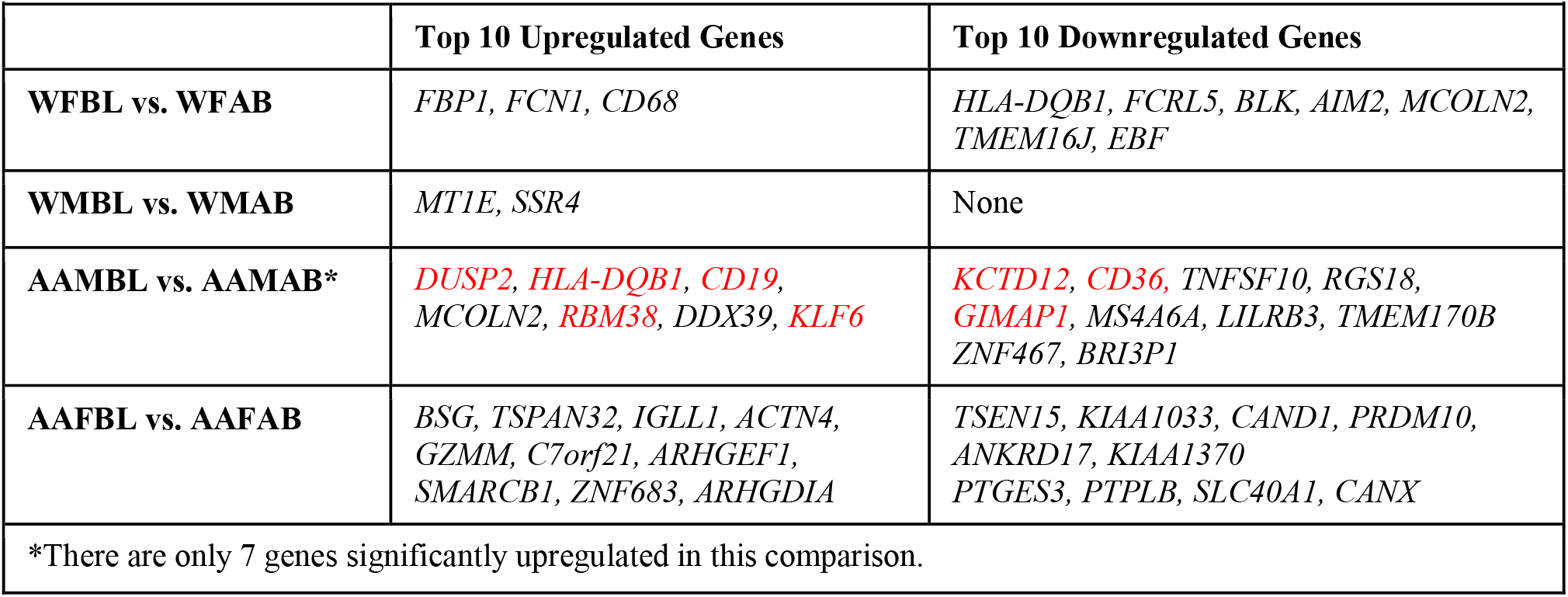
Significantly-Different Genes by Poverty Status.

**Figure 2:**
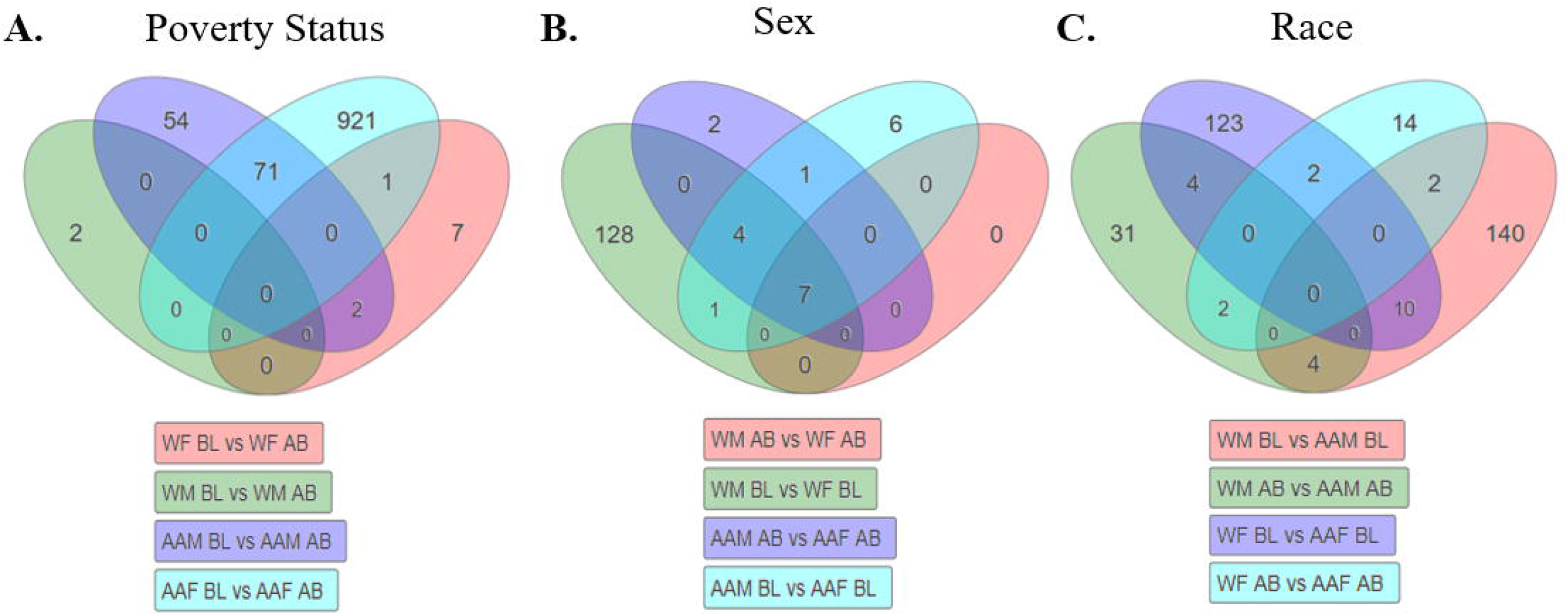
Gene expression profiles associated with poverty, sex, and race. DGE profiles of significant genes (|LogFC| >0.6, FDR AdjPvalue <0.3) in each comparison subgroup were compared by Venn diagram using iPathway Guide and stratified either by poverty status (A), Sex (B) or Race (C) to identify common and overlapping genes. N= 6/7 per group; see S1 Appendix Table 1 for completely demographics. White females below poverty (WFBL) or above poverty (WFAB); white males below poverty (WMBL) or above (WMAB); African American males below poverty (AAMBL) or above (AAMAB); African American females below poverty (AAFBL) or above (AAFAB).

We observed that few genes overall were significantly different when stratifying by sex and within the same poverty status (Figure 2B). We observed that 128 genes were unique when comparing WMs and WFs living in poverty, compared with only 6 that were significantly different between AAMs and AAFs. There were 7 genes significantly different between males and females regardless of race or poverty status (PRKY, EIF1AY, RPS4Y1, RPS4Y2, SMCY, USP9Y, and CYorf15B), of which nearly all are X- or Y-linked genes (Figure 2B). When comparing between races and within the same sex and poverty status, we identified 140 genes different between AAMs and WMs and 123 genes different between AAFs and WFs (Figure 2C). Together, we observed the most significant gene changes with respect to poverty status and this remained the primary focus of our analysis.

### Pathway Analysis of Significant Genes

We next performed a Gene Ontology (GO) analysis within iPathay to identify relevant pathways related to the significant genes differentially-expressed in individuals living in poverty. Initial GO pathway P-values were adjusted using either elimination pruning for biological processes, molecular functions, and cellular components (43) or FDR to correct for multiple comparisons for identified diseases and pathways (Figure 3). When stratified by poverty status, nearly all Biological Processes were unique to each comparative group. There were 116 biological processes related to the differentially-expressed genes in AA males living in poverty compared to those above the poverty line (Figure 3A). AA females had 91 biological processes unique to their comparative group. White males and females had fewer biological processes identified between those living in poverty vs. above the poverty line, 3 and 35, respectively (Figure 3A). There were 3 processes that overlapped between AA males and white females in poverty: positive regulation of interleukin-1 beta secretion (GO: 0050718), positive regulation of macrophage inflammatory protein 1 alpha production (GO: 0071642), and positive regulation of monocyte chemotactic protein-1 production (GO: 0071639). We observed similar patterns for each of our poverty comparisons for the GO analysis of Molecular Functions and Cellular 201 Components (Figure 3A), with few overlapping pathways between each group.

**Figure 3:**
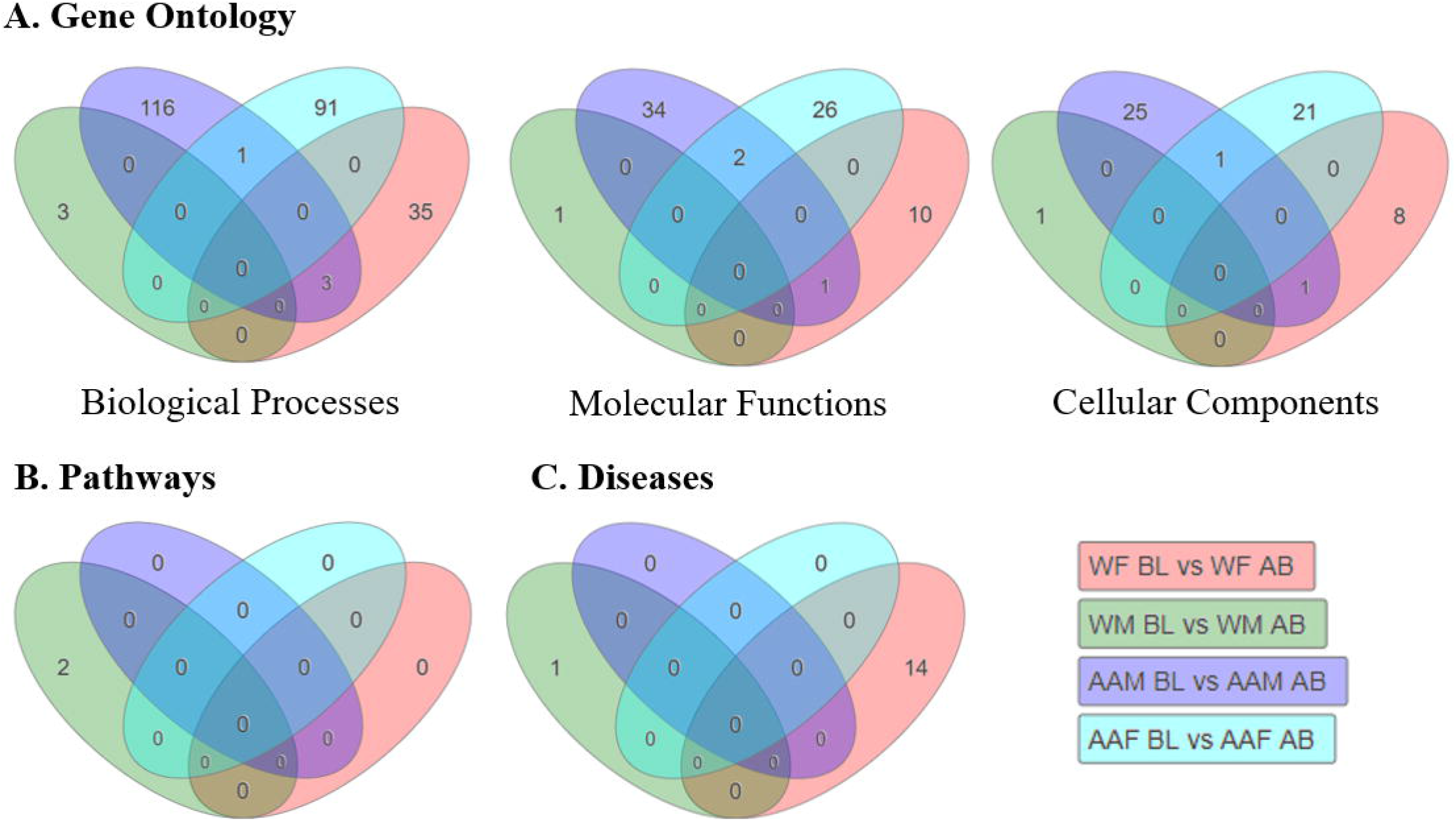
Gene ontology and pathway analysis of top poverty-related genes. Venn diagrams breaking down significant (A) GO terms related to Biological Processes, Molecular Function, or Cellular Components and stratified by poverty status for each comparison. iPathway Guide identification of unique and overlapping genetic pathways (B) or diseases (C) related to observed DGE in each comparison. N= 6/7 per group; see S1 Appendix for completely demographics. White females below poverty (WFBL) or above poverty (WFAB); white males below poverty (WMBL) or above (WMAB); African American males below poverty (AAMBL) or above (AAMAB); African American females below poverty (AAFBL) or above (AAFAB).

We next sougth to identify relevant KEGG Pathways or Diseases related to DGE in each comparative group. After adjusting for FDR, only two pathways (mineral absorption [ID: 04210] and protein processing in the endoplasmic reticulum [04141]) were significant in our entire dataset, specifically in white males in poverty (Figure 3B). We found similar results when identifying KEGG diseases (Figure 3C). One disease was significant for white males in poverty (Congenital disorders of glycosylation type I; H00118) and 14 in white females in poverty,including viral myocarditis (H00295), fructose-1, 6-biphosphate deficiency (H00114), and asthma (H00079; Figure 3C). We also conducted a GO and KEGG pathway analysis when comparing our sub cohort groups by sex and race (S1 Appendix Figure 1.) A complete list of all pathway rankings for each subgroup and comparisons in Figures 3 and S1 Appendix Figure 1 can be found in S1 Appendix.

We decided to focus the remainder of our pathway and gene expression analysis on AA males and females as both groups had the most significant gene expression differences (Figure 1 and 2) when comparing those living below the poverty line with those above in each group. GO analysis identified 1 shared biological process between AA males and females living in poverty (endosome organization; GO: 0007032; Figure 3A and 4A). There were 11 out of 22 genes within the endosome organization process that exhibited significant down-regulation in either AA males and females living in poverty (Figure 4B).

**Figure 4:**
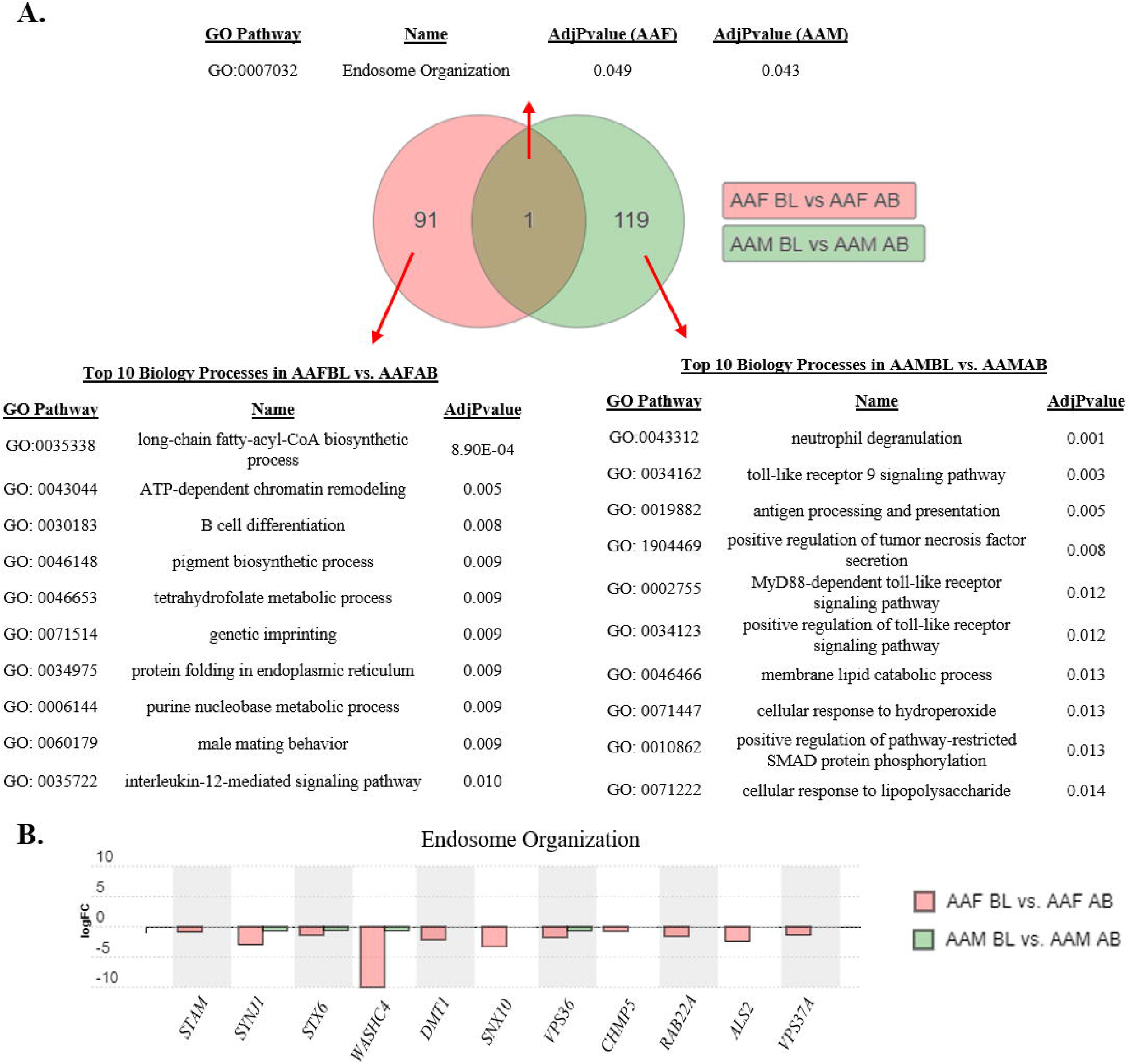
Gene ontology (GO) analysis in African American females and males. (A) Venn diagram of GO Biological Processes of significant genes identified in AAFBL vs. AAFAB compared with AAMBL vs. AAMAB. Unique and common pathways for each comparison of poverty status in females (pink) and males (green) were identified after elimination pruning P235 value adjustment to eliminate false positives. Tables provide Top 10 Biological Processes unique to AA females or AA males or overlapping between both groups. Pathways are reported with GO ascension number, name, and adjusted P-value (AdjPvalue). (B) Differential expression of significant genes identified common Endosome Organization pathway. Gene expression is reported as logFC in AAFBL vs. AAFAB (pink) or AAMBL vs. AAMAB (green).

There were 91 GO biological processes unique to AA females in poverty. Among the Top 10 most significant were long chain fatty-acyl-CoA biosynthetic processes, B-cell differentiation, genetic imprinting, and interleukin-12-mediating signaling (Figure 4A). AA males living in poverty had 119 biological processes identified in iPathway Guide compared with AA males above poverty. Top significant processes included neutrophil degranulation, toll-like receptor 9 signaling, MyD88-dependent toll-like receptor signaling, and positive regulation of toll-like receptor signaling (Figure 4A).

We also identified unique and overlapping molecular functions and cellular components in AA males and females. Two molecular functions were identified in both AA males and females living in poverty: GDP Binding (GO: 0019003) and single-stranded RNA binding (GO: 0003727; S1 Appendix Figure 2A). For both of those GO terms, seven genes were significantly down-regulated in AAs living in poverty compared with above poverty (S1 Appendix Figure 2B). Twenty-six molecular functions were unique within AA females, including top significant functions including GTPase activity and binding, nuclear receptor transcription coactivator activity, and hormone receptor binding (S1 Appendix Figure 2A). There were 35 molecular functions unique to AA males living in poverty and top pathways included Double-stranded RNA binding, insulin-like growth factor receptor binding, and toll-like receptor binding (S1 Appendix Figure 2A).

There was one cellular component GO pathway related to the early endosome (GO: 0005769) shared between AA males and females (S1 Appendix Figure 3A). Genes in this pathway were predominantly down-regulated in AAs living in poverty, particularly in AA women, where 32 of the 34 genes in this pathway were significantly down-regulated (S1 Appendix Figure 3B). Similar to our GO analysis of biological processes and molecular functions, there were more unique pathways for AA males and females. There were 21 cellular components identified as significant in AA females living in poverty, including the SWI/SNF complex, nuclear heterochromatin, and membrane rafts. Of the 26 pathways significant in AA males living in poverty, some of the most significant included phagocytic vesicle membranes,recycling of the endosome membrane, and SNARE complex (S1 Appendix Figure 3A).

### Gene Expression Validation

The expression levels of mRNAs that were identified in our microarray analysis were quantified and validated by RT-qPCR in an expanded cohort of AA males living below (n=27) or 273 above (n=29) poverty (S1 Appendix). We chose to validate genes in AA males because this demographic group is at the greatest risk for mortality in Baltimore City (24). We selected genes identified in our microarray analysis within the top 10 up-regulated or down-regulated between AA males living in poverty compared with those living above poverty (Table 1, red), as well as non-significant control genes and genes of interest related to immune signaling or inflammation.

We validated that there is a significant difference in the expression of KLF6, DUSP2, RBM38, CD19, and KCTD12 between AA males living in poverty versus those living above poverty (Figure 5. With the exception of KCTD12, the rest of the genes had lower levels of expression in AA males living in poverty. We were unable to validate the expression levels of CD36, HLA-DQB1, or GIMAP1, which were significant in our microarray but not our expanded cohort (Figure 5). Collectively, these data indicate that our microarray analysis and validation approach can identify genes differentially-expressed in populations living in poverty.

**Figure 5:**
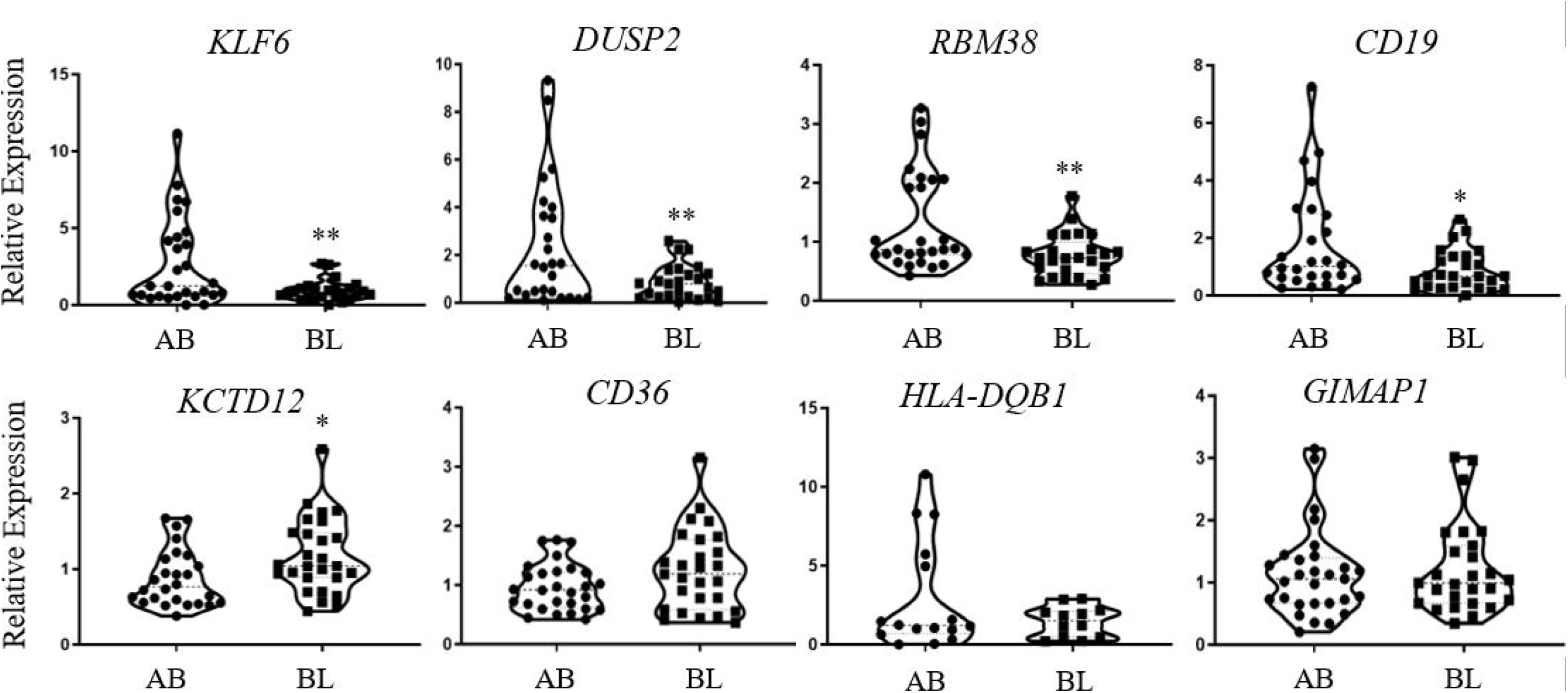
mRNA validation in African American males living in poverty. 11 mRNAs identified from our microarray analysis were validated using real-time quantitative PCR in African American males living above (AB) or below (BL) poverty. Genes were quantified using mRNA-specific primers and outliers were excluded. All genes were normalized to the combined average of ACTB and GAPDH. Violin plots of the eleven genes compared BL (n=27) vs AB (n=29). *P<0.05, **P<0.01; Student’s t-test.

## Discussion

Our analysis identified several significantly and differentially-expressed mRNAs in PBMCs associated with poverty status in Baltimore City residents. AA males and females living in poverty had more genes differentially-expressed than whites when compared with their above poverty controls. In AA males, we used RT-qPCR to validate the expression of several of the transcripts identified in our microarray within an expanded cohort (Figure 5). Unexpectedly,several significant genes, including KLF6, DUSP2, RBM38, and CD19, were found to be down-regulated in AA males living in poverty, opposite of our initial findings in our microarray. This may be due to the fact that our microarray was performed in six or 7 individuals, as well as the fact that RT-qPCR is much more accurate in single-gene expression validation compared with global discovery analysis (44). We did not identify any transcripts to be significant in our RT-qPCR analysis that were not identified in our microarray.

The low levels of expression of these transcripts in AA males living in poverty offers several intriguing paths to pursue during follow-up analyses. Kruppel-like factor 6 (KLF6) has well-defined roles as a transcriptional activator in endothelial cells and regulates the repair of vascular damage after tissue injury (45). KLF6 also represses the expression of B-cell leukemia 6 (BLC6) in macrophages and monocytes to promote pro-inflammatory gene expression patterns necessary for alleviating tissue damage. When KLF6 was repressed in mouse models, the required inflammatory response from macrophages to promote healing was attenuated (46, 47).This response network is also mediated through miR-223 and hypoxia inducible factor 1 alpha (HIF1A), both of which are important node regulators of vascular inflammatory response (48, 49). RNA-binding motif protein 38 (RBM38) has identified roles in endothelial repair following arterial injury (50) and deficient mouse models develop an accelerated aging phenotype and have impaired hematopoiesis (51).

We also identified low levels of dual specificity phosphatase 2 (DUSP2) in AA males living in poverty. DUSP2 negatively regulates ERK1 and ERK2 signaling within the MAP kinase signaling pathway and is important for T-cell gene expression regulation (52). DUSP2 inhibition in mice leads to susceptibility to colitis and inflammation and increase secretion of cytokines from T_H_17 cells (53). If KLF6, RBM28, or DUSP2 may be novel mechanisms in which susceptibly to vascular injury or inflammatory-related conditions is promoted in AA males living in poverty. This work could provide some key explanations of how inflammation and stress is linked to social disadvantage. Futures studies experimentally validating this hypothesis are warranted.

Our GO analysis of gene expression in AA males identified several pathways related to TLRs, toll-like receptor 9 (TLR9) signaling, MyD88-dependent TLR signaling, and positive regulation of TLR signaling, with potential functions relevant to living in poverty and driven by lower expression levels of related TLR1 and TLR8 (Figure 4A). Most TLRs are bound within the membrane of the endosome, including TLR8, and several are found in the plasma membrane of the cell, including TLR1. The TRL family of enzymes are heavily involved in the activation of innate immunity within white and peripheral blood cells, inflammation response, as well as playing a variety of roles related to adaptive immunity (54). A recent analysis of AA whole-blood transcriptomes identified low SES association with up-regulation of TRL1 and TRL8 (34).We found TLRs to be down-regulated with poverty in our cohort, highlighting the need to further study these genes and related pathways in response to social and environmental stress within those living in impoverished conditions over a long period of time. It is possible adverse conditions affect different communities in different ways on the level of a single gene, yet have similar consequences when looking across multiple genes within the same pathway.

AA women living below poverty had the most genes differentially-expressed compared with above poverty controls (Figure 1). There is significant down-regulation of genes related to fatty-acid biosynthesis and metabolism, including PTPLB, ACSL3, and ACSL5, and transcriptional regulators including HDAC2, HDAC4, and SP3. HDAC2 and HDAC4 are part of the family of histone deacetylases and prior studies have reported that each have been linked with vascular inflammation and Angiotensin II-induced autophagy (55, 56). These genes have not been associated with poverty status before.

Our gene enrichment analysis identified endosome organization (GO: 0007032) as the only biological process significant in both AA males and females living in poverty. The endosomal compartments within immune cells are critical for extracellular signaling response and for appropriate activation of inflammatory responses to external stimuli (57). Endosomal trafficking of several TLRs is important for this response (58) and perturbations in TLR activation or in dysfunctional endosomes can lead to autoimmune conditions and an increase in cellular inflammatory response resulting in downstream inflammation (58–60). While there have been no prior studies associating endosomal functioning with SES, behavioral work in macaques identified social status-related differences in toll-like receptor 4 (TLR4)-mediated inflammation via downstream transcriptional NF-kB activation (61).

Prior studies have linked gene expression in PBMCs with poverty status in HANDLS participants. For example, profiling of young and old AAs and white males living above or below poverty identified hundreds of mRNAs and long, non-coding RNAs associated with poverty in white males. This included enrichment for genes related to stress and immune signaling in whites living above poverty (62). Poverty status and social adversity are also associated with differential expression of stress- and inflammatory-related genes within the conserved transcriptional response to adversity (CRTA) network (32, 34–36). There are 53 genes in the CRTA network including cytokines, inflammatory transcriptional regulators (including NF-kB), and members of the Type I interferon response. Their expression is susceptible to environmental stressors (63). When comparing the CRTA gene set with our microarray analysis,we did find some overlap in significant genes. AA males living in poverty have significantly lower levels of interferon induced with helicase C domain 1 (IFIH1), a member of the Type I interferon response pathway. AA females in poverty also expressed IFIH1 at significantly lower levels, in addition to lower levels of inflammatory transcript factor NFKB1, and higher levels of preB-cell receptor immunoglobulin lambda like polypeptide 1 (IGLL1) compared with AA females living above poverty (data not shown). We did not observe any overlap with the CRTA network and expression differences in whites in our study. Together, this suggests additional studies are needed for these over-lapping transcripts, as these CRTA-related genes have been identified in several independent cohorts.

Our and other related analyses examining gene expression profiles related to poverty status highlight an important question: how does environmental stress mechanistically induce these changes? Prior analysis of DGE in AA and white women with hypertension identified differentially-expressed microRNA (miRNA) profiles associated with differentially-expressed mRNAs in AA hypertensives (64, 65). miRNAs post-transcriptionally regulate gene expression levels and aberrant expression of these regulatory RNAs can lead to more aggressive diseases, including cancer in AAs (66, 67). We used TargetScan’s default settings (68) to predict and identify possible miRNA regulators of each of the top genes identified in our study in AA males and females (miRNAs listed in S2 Appendix). In vitro confirmation of these results is necessary.

Allelic variation and methylation can also influence gene expression levels. Variants likely to promote up-regulation of proinflammatory cytokine genes IL-1 and IL-6 and down-regulation of anti-inflammatory IL-10 are more likely found in AA women (69). Ancestral alleles can strongly influence expression quantitative trait loci (eQTLs) governing stress- and pathogen-response gene expression in immune cells (70). Recent studies also suggest that neighborhood disadvantage and SES at birth are correlated with differential CpG methylation states (71, 72). Increased methylation states of inflammatory genes in individuals coming from SES disadvantage have been linked with differential expression of FKBP5, CD1D, F8, KLRG1, and NLRP12 (73). Understanding environmental stress response by untangling the connections between allelic variation, epigenetic change, and pre- and post-transcriptional regulation will pinpoint the most important pathways that contribute to gene expression influenced by living in poverty. This may then offer new strategies to prevent these changes from causing downstream health complications in AA males and females and others who live in poverty.

There are a few limitations to this study. While the HANDLS study is a representative sampling of residents in Baltimore City, gene expression and pathway validation in additional independent diverse cohorts is warranted to identify if the pathways we have identified here are representative within AAs living in other parts of the United States. Our study was not able to discern which factor of living within poverty is primarily associated with DGE in our cohort, for example food insecurity or nutritional factors, the influence of psychosocial stress or perceived violence, or many of the other complex environmental influences resulting from living in an urban impoverished area. This initial study also does not identify a specific biological mechanism through which poverty transduces physiological responses. It’s possible these predispositions are channeled through just one or several key genes unique to each AA and white males and females or that the few overlapping genes identified here are the most relevant. This analysis was a cross-sectional study of gene expression in HANDLS. A longitudinal analysis may provide deeper insight into the most important pathways and genes affected by living in poverty.

We analyzed gene expression within a mixed population of peripheral blood mononuclear cells and did not discern whether DGE profiles were associated with unique cell types. Follow-up analyses are warranted to experimentally validate these initial findings and identify if DGE profiles in specific cell types serve as better biological biomarkers for the effect of poverty status on gene expression. Results from our initial analysis can be validated with such an approach as well as inform targeted gene arrays or RT-qPCR validation studies.

In conclusion, our study provides additional evidence that gene expression patterns in PBMCs are associated with poverty status, particularly in African Americans. We observed that genes related to endosomal function and toll-like receptor signaling are differentially-expressed in AA males living in poverty. Impoverished environments have long been associated with an elevation in prevalance of health disparities. This work helps elucidate the genetic pathways potentially suspectible to environmental stress in AA males and females. While these findings are promising, a more nuanced examination of what is it about poverty status that influences governs these gene expression differences is needed. Disentangling other factors that are often associated with poverty status such as segregation, high crime, low quality housing, and poor education will be useful for successful interventions. Together, this deeper understanding of the social determinants of health on gene expression can provide new avenues for disease prevention and intervention.

## Materials and Methods

### Cohort Demographics

Gene expression analysis was performed on peripheral blood 433 mononuclear cells (PBMCs) isolated from a subcohort of participants in the Healthy Aging in Neighborhoods of Diversity Across the Life Span (HANDLS) study of Baltimore City residents(28). HANDLS is a longitudinal, epidemiological study of a diverse and representative sample of Baltimore City residents and was designed and implemented by the National Institute on Aging Intramural Research Program (NIA IRP) (28). The subgroups for this study included age-matched African American (AA) or white (W), males (M) or females (F) who were living above (AB) or below (BL) 125% of the 2004 federal poverty guideline in Baltimore City at the time of collection (n=6-7/group; S1 Appendix for complete demographics). All blood samples were collected in the morning after an overnight fast. The isolation of the PBMCs was conducted within 3 hours of blood draw, aliquoted, and stored at −80°C. The Institutional Review Boards of the National Institute of Environmental Health Sciences and Morgan State University approved this study and a written informed consent was signed by all participants.

### RNA isolation and Microarray

Total RNA was isolated from PBMCs using Trizol (Invitrogen)phenol/chloroform and subjected to DNase digest. Total RNA quality and quantity was assessed with a Nanodrop 2000 and an Agilent Bioanalyzer. Global gene expression in PBMCs was analyzed via microarray using the Illumina Beadchip HT-12 v4 (San Diego, CA). Total RNA was prepared according to the manufacturer’s protocol and raw signal data was filtered as previously described (64, 65). Briefly, probe signal data were subjected to Z-score and FDR normalization to identify outliers. Individual genes with pair-wise z-test P-values < 0.05, average intensity >0, |Z-ratios| > 01.5-fold, and FDR ≤0.3 were considered significant and were based on the maximum score identified from multiple probes of the same gene. The logFC and FDR values were next imported into iPathway for further analysis (see below). The microarray data has been submitted to the Gene Expression Omnibus (GSE149256).

### Gene and Pathway Analysis

We uploaded our microarray gene sets into Advaita iPathwayGuide(Ann Arbor, MI) to identify the top up- and down-regulated genes that were poverty-related (39–42). Each gene set for AA males, AA females, white males, and white females compared those living below 125% of the 2004 federal poverty line at the time of sample collection with those living above. Genes were considered significant if |LogFC| >0.6 and FDR AdjPvalue < 0.3 to reduce false positives and plotted on volcano plots using iPathway. Our pathway enrichment analysis was complied with the gene ontology (GO) tool of iPathway Guide Program. Gene ontology terms were biological process, molecular function, and cellular components associated with significantly differentially-expressed genes. A P-value correction was applied with an elimination pruning (43) to remove potential false positives for the GO analysis. False positives for the biological and disease pathways tools in iPathwayGuiede were removed with a p-value correction using FDR.

### RT-qPCR

Total RNA from AA males living above or below the poverty line was reverse transcribed into cDNA using SuperScript II reverse transcriptase (Invitrogen) and random hexamers. Levels of each target transcript were quantified by RT-qPCR using 2x SYBR green PCR master mix (ThermoFisher) and gene-specific primers (see S1 Appendix for forward and reverse sequences). All reactions were run in duplicate using an Applied Biosystems QuantStudio 6 Flex System Real-Time using the 384-well block (Thermofisher). Transcript expression levels were normalized to the combined average of GAPDH and ACTB and compared between AA men living below or above the poverty line using the 2^-ΔΔt^ method (74).

### Statistical Analysis

A one-way ANOVA was used when comparing multiple group means for each physiological metric in our cohort and a Fisher’s Exact Test was used to determine the the association between poverty status and disease history or smoking status. mRNA expression levels in the validation cohort were examined for Gaussian distribution by measuring skewness, kurtosis, and using a visual inspection of histograms. Outliers from each group for each validated were removed using Grubb’s Test with an α = 0.05. Violin plots were generated using GraphPad Prism 8 software. A Student’s t-test was used when comparing two groups, unless indicated otherwise. A P-value of <0.05 was considered statistically significant unless otherwise specified.

## Data Availability

The microarray data has been submitted to the Gene Expression Omnibus (GSE149256).

## Acknowledgments

The authors thank Althaf Lohani and David Freeman for their technical assistance and Dr. Kevin Becker for his input. We thank Dr. Ngozi Ejiogu and the HANDLS staff for the management and careful evaluation of study participants.We thank Dr. Cordelia Ziraldo for her assistance and training using iPathwayGuide software.

